# A Web-based Spatial Decision Support System of Wastewater Surveillance for COVID-19 Monitoring: A Case Study of a University Campus

**DOI:** 10.1101/2021.12.29.21268516

**Authors:** Wenwu Tang, Tianyang Chen, Zachery Slocum, Yu Lan, Eric Delmelle, Don Chen, Neha Mittal, Jacelyn Rice-Boayue, Tarini Shukla, Sophia Lin, Srinivas Akella, Jessica Schlueter, Mariya Munir, Cynthia Gibas

## Abstract

The ongoing COVID-19 pandemic has produced substantial impacts on our society. Wastewater surveillance has increasingly been introduced to support the monitoring, and thus mitigation, of COVID-19 outbreaks and transmission. Monitoring of buildings and sub-sewershed areas via a wastewater surveillance approach has been a cost-effective strategy for mass testing of residents in congregate living situations such as universities. A series of spatial and spatiotemporal data are involved with wastewater surveillance, and these data must be interpreted and integrated with other information to better serve as guidance on response to a positive wastewater signal. The management and analysis of these data poses a significant challenge, in particular, for the need of supporting timely decision making. In this study, we present a web-based spatial decision support system framework to address this challenge. Our study area is the main campus of the University of North Carolina at Charlotte. We develop a spatiotemporal data model that facilitates the management of space-time data related to wastewater surveillance. We use spatiotemporal analysis and modeling to discover spatio-temporal patterns of COVID-19 virus abundance at wastewater collection sites that may not be readily apparent in wastewater data as they are routinely collected. Web-based GIS dashboards are implemented to support the automatic update and sharing of wastewater testing results. Our web-based SDSS framework enables the efficient and automated management, analytics, and sharing of spatiotemporal data of wastewater testing results for our study area. This framework provides substantial support for informing critical decisions or guidelines for the prevention of COVID-19 outbreak and the mitigation of virus transmission on campus.

## 1. Introduction

The COVID-19 pandemic has fueled a renewed interest in wastewater-based epidemiology. Wastewater testing for traces of viral and bacterial pathogens has been used for decades to track and manage outbreaks of infectious disease (Prado et al., 2012; Tambini et al., 1993). Early reports in mid-2020 demonstrated that wastewater concentrations of SARS-CoV-2 could serve as a leading indicator for cases detected by clinical testing within city sewersheds (Ahmed et al., 2021; Peccia et al., 2020), with collection of samples from wastewater treatment plant influent providing coverage of entire cities or large neighborhoods. The practical application of monitoring at city scale is primarily to detect infection trends in communities, which has been especially useful in the case of COVID-19, both because COVID-19 infections may be asymptomatic for several days prior to detection of cases by testing, and because especially in the early months of the pandemic, testing capacity lagged behind the rapid spread of the disease. In such scenarios, wastewater testing can serve as a leading indicator of the increase of disease incidence in an urban area. There has also been an increasing interest in monitoring in neighborhood or smaller scale areas for the presence of the SARS-CoV-2 virus in wastewater, because such small-scale monitoring can provide evidence to support targeted public health interventions including distribution of masks or selection of populations for increased testing (Bowes et al., 2021).

COVID-19 is easily transmitted in congregate living situations, with early and devastating outbreaks being documented in nursing homes and jails (Kırbıyık et al., 2020; Lam-Hine et al., 2021). Beside these, other indoor settings such as schools (including universities), restaurants, and hospitals have been identified as having high risk for the spread of COVID-19 (Fox et al., 2021; Lam-Hine et al., 2021). Many universities attempted to implement some type of wastewater surveillance program during the early months of the pandemic, with varying degrees of success (Gibas et al., 2021; Harris-Lovett et al., 2021; Karthikeyan et al., 2021). To effectively detect and monitor outbreaks of COVID-19 in these indoor settings requires wastewater surveillance capabilities at small spatial scales such as building level. The study reported in this article is focused on building-level wastewater surveillance for COVID-19 monitoring from a spatiotemporally explicit perspective.

Wastewater surveillance typically requires a set of sequential steps, including sample site setup, sample collection (including storage and shipping; per CDC Wastewater Surveillance strategy), lab analysis, and subsequent analysis and visualization of wastewater testing results and associated data. Geographic Information Systems (GIS) methods have been applied for the management and mapping of spatially explicit data related to wastewater testing and COVID-19 monitoring, and dashboard techniques have gained increasing attention due to their visual presentation capabilities within web-based environments (Dong et al., 2020; Lan et al., 2021; Naughton et al., 2021). Yet, most of the existing dashboards for COVID-19 and wastewater studies only concentrate on management and visualization of relevant spatial or spatiotemporal data; their support on spatial analytics and modeling capabilities is inadequate. Spatial analytics and modeling, however, are pivotal in discovering patterns of interest hidden in complicated spatiotemporal data, and providing predictive or scenario analysis capabilities for monitoring and mitigation of pandemic situations (Franch-Pardo et al., 2020). Spatial Decision Support Systems (SDSS) hold potential in filling this gap.

SDSS, which originated from the domain of Geographic Information Science (Armstrong et al., 1986; Sugumaran & Degroote, 2010), have been increasingly applied to assist with decision making within spatially explicit contexts. SDSS is based on (but more than) the integration of decision support systems and GIS, and provides inherent support for spatial analytics and modeling capabilities. This makes SDSS unique and powerful in informing decision making processes associated with complex spatial or spatiotemporal phenomena. A variety of applications such as environmental monitoring, natural resources, public health, transportation, and land use and land cover change have built SDSS to address complex decision problems within spatially explicit contexts (Delmelle et al., 2014; Keenan & Jankowski, 2019; Sugumaran & Degroote, 2010). In particular, driven heavily by Internet technologies and cyberinfrastructure (NSF, 2007), web-based SDSS has received much attention over the past few years (Lan et al., 2020; Lee et al., 2017; Tayyebi et al., 2016). While a growing body of the literature has highlighted the power of web-based SDSS, the applications of web-based SDSS for the resolution of complex spatiotemporal decision problems in general and small-scale wastewater surveillance for COVID-19 monitoring, in particular, remain scant.

In this article, we describe a web-based SDSS framework for building-level wastewater surveillance. We used a university campus (the main campus of the University of North Carolina at Charlotte) as a study case. This framework supports the automated synchronization and update of lab test results, space-time cluster analysis for identifying hotspots of COVID-19 incidents at the building level over time, and automated update of dashboards within web-based environments. The integration of these geospatial data and analytics capabilities play a critical role in providing timely information on COVID-19 incidents in the study region over time.

Specifically, we focus on addressing the following sets of research questions in this study: 1) Are there any space-time clusters of positive wastewater testing results at the building level and where are they? 2) What are those sampling sites that exhibit similar responses over time in terms of wastewater testing results and where are they?

The remainder of this article is organized using the following structure. In section 2, we discuss the background and relevant literature of this study. In section 3, we present the study area and data, the design of the entire web-based SDSS framework as well as its implementation. Section 4 presents the results including space-time cluster analysis, and Section 5 gives relevant discussion. Section 6 concludes this article.

## 2. Literature Review

### 2.1. Wastewater Surveillance

A typical workflow for building-level wastewater surveillance includes collection of a sample at regular intervals with laboratory results within 24 hours of collection. Samples can be collected using a variety of methods (Medema et al., 2020), ranging from collection of a sample volume at one timepoint (a “grab” sample), to composites collected by passive sampling for example using fibrous swabs (Liu et al., 2021), and composites collected using pump autosamplers which add to the sample at regular intervals over the course of a day prior to collection. Once collected, samples are processed and concentrated. A wide variety of methods are available for this concentration step as well, and choice of method is governed by a combination of viral recovery efficacy, cost, materials availability, and processing time, as described in our previous work (Juel et al., 2021). RNA is extracted from the concentrated sample, and virus is quantified using a molecular detection protocol such as RT-qPCR or RT-ddPCR (Barua et al., 2021; Ciesielski et al., 2021), which provides a viral concentration in terms of copies of virus per liter of wastewater collected. This value can be used effectively as a simple binary indicator of positivity, as demonstrated in the pilot phase of our campus monitoring program (Gibas et al., 2021) but also has the potential to connect the information about population size and volume of water used in the building to provide an estimate of the number of individuals who might be SARS-CoV-2 positive (Sweetapple et al., 2022). Once a positive signal is detected, a decision is made about whether to test all individuals in that building, after consulting institutional information about individuals who have recently tested positive or been connected to that site via contact tracing. If there are no previously-known individuals who are likely to be the source of the positive signal, then the entire building population is subjected to clinical testing.

While many institutions and localities have deployed wastewater testing for SARS-CoV-2 during the pandemic, only a small fraction of these projects have so far made data available in service of larger efforts to develop quantitative models and consistent practices in wastewater epidemiology (Naughton et al., 2021). Data dashboards are a common means for sharing such information when it is made available, and in some cases have been incorporated into state-level public health reporting (e.g., see https://covid19.ncdhhs.gov/dashboard/wastewater-monitoring). Dashboard techniques have been extensively applied for the sharing of data related to COVID-19. A number of dashboards have been developed and deployed to support the wastewater surveillance initiatives for the monitoring of COVID-19 worldwide. For example, there are a number of dashboards registered via the web site of COVIDPoops19 project (Naughton et al., 2021). About 40% of these dashboards have built-in Web GIS functionality. The software platforms used to present these dashboards include Esri ArcGIS Online, Tableau, R Shiny, Microsoft Power BI, and CARTO. The first three (ArcGIS Online, Tableau, and R Shiny) are the dominant choices for the implementation of wastewater surveillance dashboards. Most of the wastewater data managed and reported by these wastewater dashboards are at the wastewater treatment plant level and collected weekly, while a smaller number of projects report daily or multiple days per week. A few universities make campus wastewater data available in real time via public dashboards (e.g. University of California at San Diego, Clemson University), but in other cases, for instance at the University of North Carolina at Charlotte, the concern of upper administration not to alarm students or parents with details of wastewater alerts has resulted in a decision to keep this information for internal use only. A number of existing dashboards only focus on the visual presentation (in maps or charts) of wastewater-related data, and may not provide the spatiotemporal analytics and modeling of wastewater testing results and relevant data. The need for spatiotemporal analysis and modeling to guide the study of wastewater testing results for the monitoring of COVID-19 outbreak and prevention has been recognized in the literature (Karthikeyan et al., 2021).

### 2.2. Spatial Decision Support Systems

SDSS are integrative computer-based systems that provide decision-making support for complex spatial problems via the fusion of spatial data management, modeling, and visualization capabilities (Densham, 1991; Malczewski, 1999; Sugumaran & Degroote, 2010). SDSS, with an origin from Decision Support Systems (Marakas, 2003), are distinguished by their ability to handle decision-making support within a spatially explicit context via the incorporation of GIS-based functionality. Yet, SDSS differ from GIS in that they encompass spatial modeling capabilities to aid decision-making (Armstrong et al., 1986; Sugumaran & Degroote, 2010). For example, with the incorporation of a spatial simulation model, SDSS can enable what-if scenario analysis to explore potential alternative solutions of a spatial problem. The spatial optimization model helps SDSS identify spatially explicit optimal solutions facing decision makers (represented by site selection problems). Further, spatial statistical models allow for the discovery of spatial patterns of interest (e.g., clusters of disease or accidents) from spatial data. All these modeling capabilities can be built within a SDSS that informs and facilitates decision making processes associated with complex spatial or spatiotemporal problems (Ghosh, 2008). In terms of implementation, a SDSS includes the following functional modules: data management, model management, visualization and report generation, and a user interface (Armstrong et al., 1986; Densham, 1991; Sugumaran & Degroote, 2010).

While the study of SDSS in early stages focuses on the development of conceptual architecture, cyberinfrastructure-enabled computing technologies such as web and cloud computing have been fostering the implementation and applications of SDSS into different domain studies (Sugumaran & Degroote, 2010; Tang et al., 2017). For example, Mwaura and Kada (2017) presented a web-based SDSS in which a multi-criteria decision making model was used to evaluate potential sites of geothermal wells in Kenya, east Africa. Crimi et al. (2019) investigated the identification of priority regions in Bradford, UK for freight lorry parking within a web-based SDSS environment. Lan et al. (2020) applied web-based SDSS that guides the monitoring and sharing of water quality information of private wells in Gaston County, NC, USA. Spatial interpolation algorithms were used in Lan et al.’s work to generate the spatially continuous distribution of water quality that will inform residents or governments for potential water contamination.

## 3. Materials and Methods

### 3.1. Study Area and Data

Our study area is the main campus of the University of North Carolina at Charlotte, USA (see Fig. 1). The main campus of the University (35°18’25”N, 80°44’06”W) is located in the north of the City of Charlotte (within Mecklenburg County). The University is an urban university with about 3,000 employees (including faculty and staff); and 30,146 students in the Fall semester of 2020. Among them, around 6,000 students are living in residential halls on campus. In total, there are 138 buildings in the main campus, 33 residence halls, 32 academic buildings, and 73 other types. Please see Appendix 1 for sources of the aforementioned information about the University. In terms of topography, the main campus is high in east and west and low in the middle (range of elevation: 176-226 meters). The slope of the main campus varies from 0° to 25° (based on a 1-m DEM derived from LiDAR point cloud data). The Toby Creek area is the lowest-lying region on campus. Toby Creek flows through the campus and discharges into Mallard Creek at the north end of the campus. The university’s sewer system is composed of gravity sewer lines, where a sampling at a specific sewer manhole location will be affected by upstream nodes. Lateral and branch sewer lines collect wastewater from all residence and academic buildings, and then connect to a main sewer line (Charlotte Water’s wastewater system) which parallels Toby Creek. Campus wastewater is treated at the nearby Mallard Creek Treatment Facility.

**Fig. 1.**
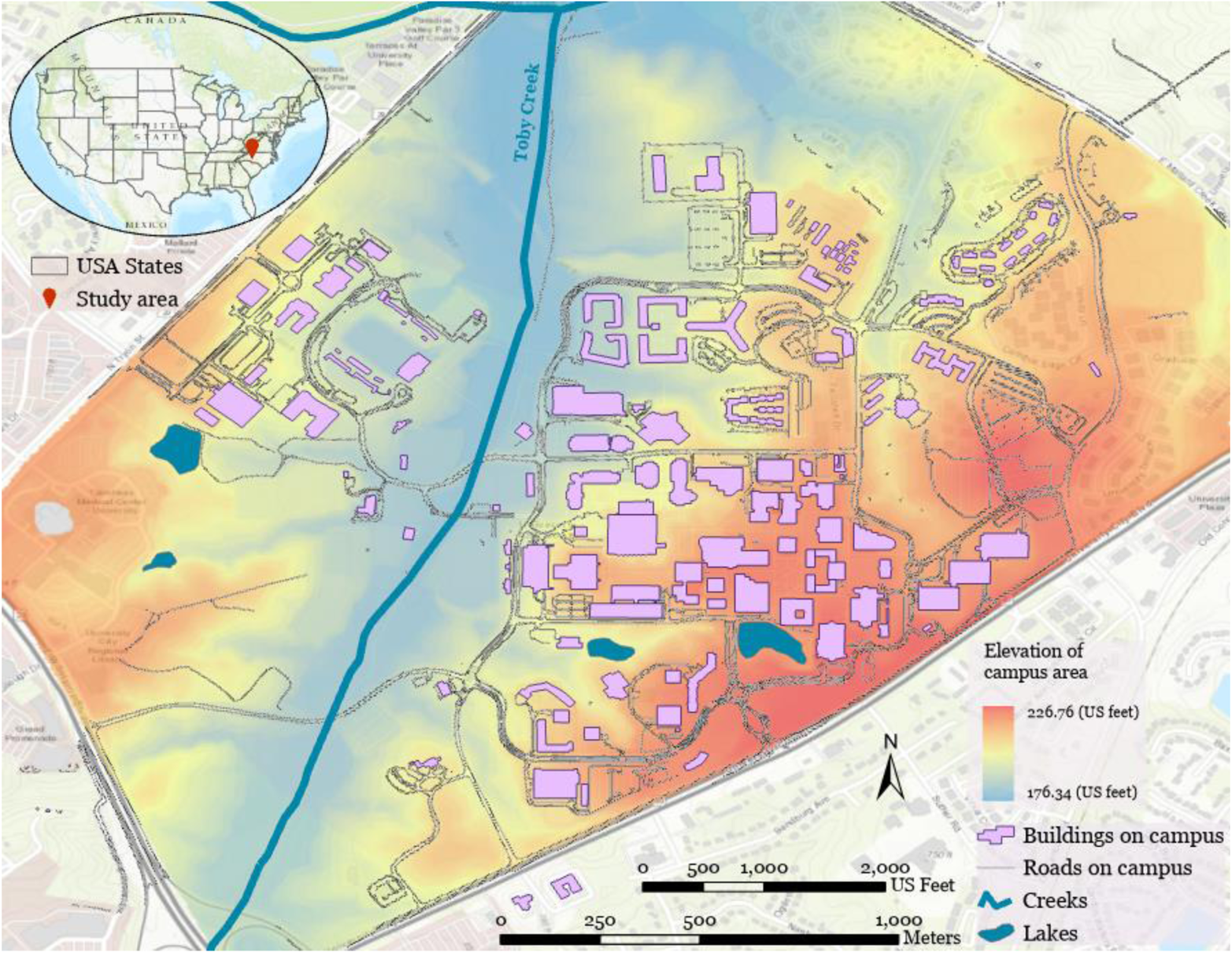
Map of the main campus of the University of North Carolina at Charlotte, USA (sewage network details are not shown for the protection of physical security of university infrastructure).

The University of North Carolina at Charlotte launched its wastewater-based epidemiology (WBE) surveillance program in late Summer 2020 to assist the University in monitoring COVID-19 incidence. Wastewater signal has been used since that time to identify dormitory populations for testing (“surge testing”) in the event of detection of SARS-CoV-2 virus in the absence of a previously identified source. The wastewater surveillance program has been collecting and analyzing wastewater samples since September, 2020. A team of faculty, staff, and students from bioinformatics, engineering, computer science, and geography collaborate to develop this monitoring system, with infrastructure support from the University’s Facilities Management staff. The WBE team has also developed a Building Information Modeling (BIM; see Becerik-Gerber et al. (2012)) 3D model for each residence hall on campus. Each BIM model includes the building envelope and plumbing fixtures, which can be used to identify rooms and zones in which potential infected individuals are located. Wastewater data collected together with BIM models have allowed campus administration to make timely and targeted decisions to prevent the cluster outbreak and spread of COVID-19 on campus (see Gibas et al., 2021 for detail). We collected spatial data to support the wastewater surveillance work for our study area. These data include buildings, sewer lines, sampling sites, road network, and elevation.

There are in total 38 sampling sites that were identified and established for wastewater collection since Fall 2020 (see Fig. 2 for illustration). These sampling sites are organized in two types: for residence halls (a sampling site covers a building or part of the building) and for buildings within a sub-sewershed—referred to as neighborhood site in this study (a sampling site covers multiple buildings). Manholes and plumbing cleanouts are selected to set up these sampling sites. As a manhole may connect to multiple sewage lines from different buildings, a manhole may have multiple auto-samplers with probes deployed in different directions (up to two in our study) installed to collect sewage samples from different buildings. Further, a building (typically large) may have two or more sampling sites each covering different parts of the building. These sampling sites cover in total 89 buildings on campus for wastewater monitoring. We used a Trimble GPS handheld unit (with a submeter accuracy) to obtain the coordinates of the sampling sites. However, 10 of 38 samplers are located either very close to the building or inside the building, which degrades the signal quality of GPS satellites. Therefore, their locations are determined using Google Earth and images taken using a digital camera. One sampling site is completely under trees with dense canopy, where we cannot determine its exact coordinates using a GPS instrument or Google Earth imagery. In such a case, we used the location of the corresponding manhole (identified from the GIS data of the sewage network) as the coordinates of the sampling site.

**Fig. 2.**
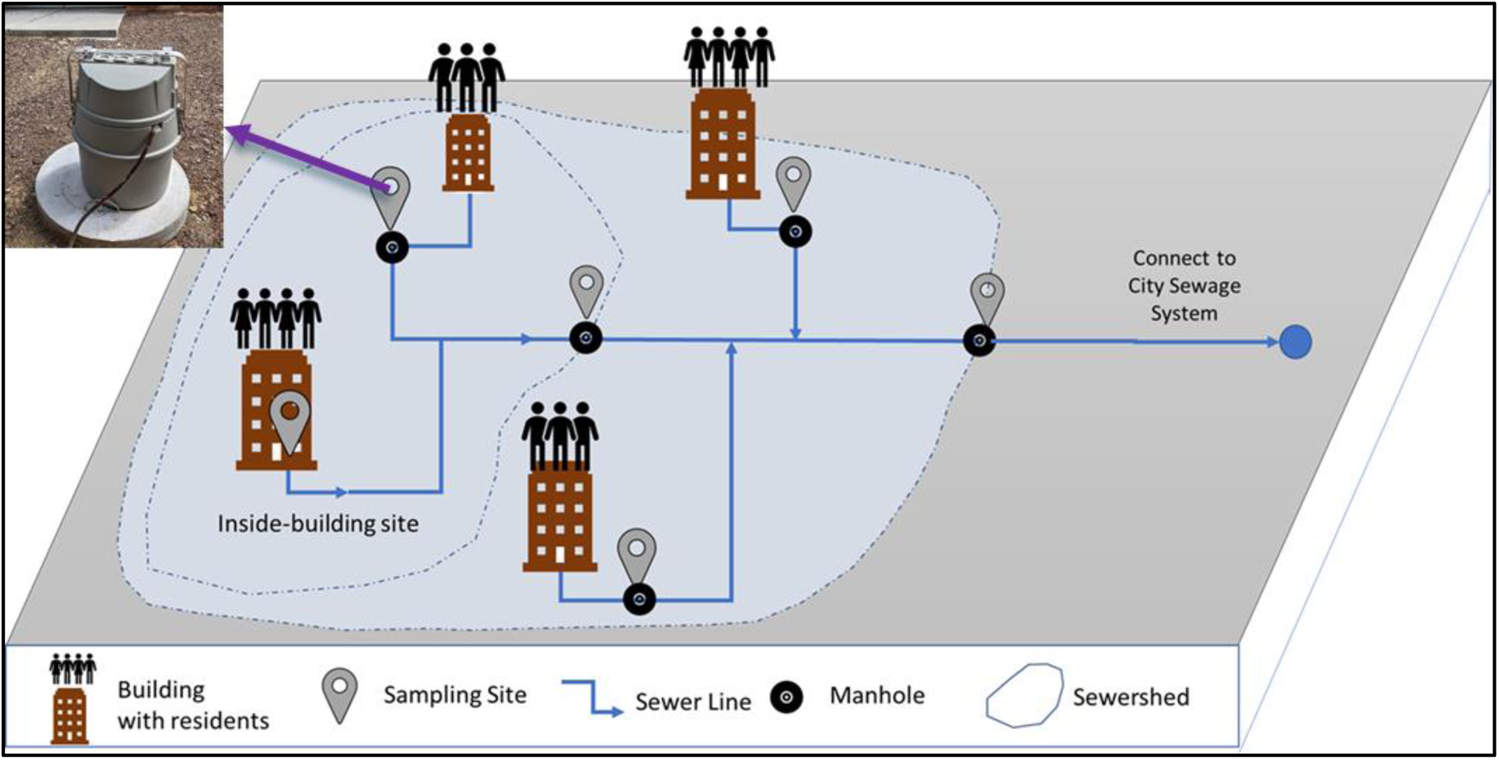
Illustration of sampling site setup for building-level wastewater surveillance.

### 3.2. Methods

In this section, we present the framework of the web-based SDSS and its main components. Fig. 3 illustrates the design of the web-based SDSS framework for wastewater surveillance in this study. This framework supports the data management, model management, and visualization of wastewater data that are spatiotemporally explicit. The integration of these functionality allows for the automated synchronization of wastewater testing results, on-demand spatiotemporal analysis of COVID-19 incidents from wastewater results, and automatic update of Web GIS dashboard that supports timely decision making in a spatially explicit manner.

**Fig. 3.**
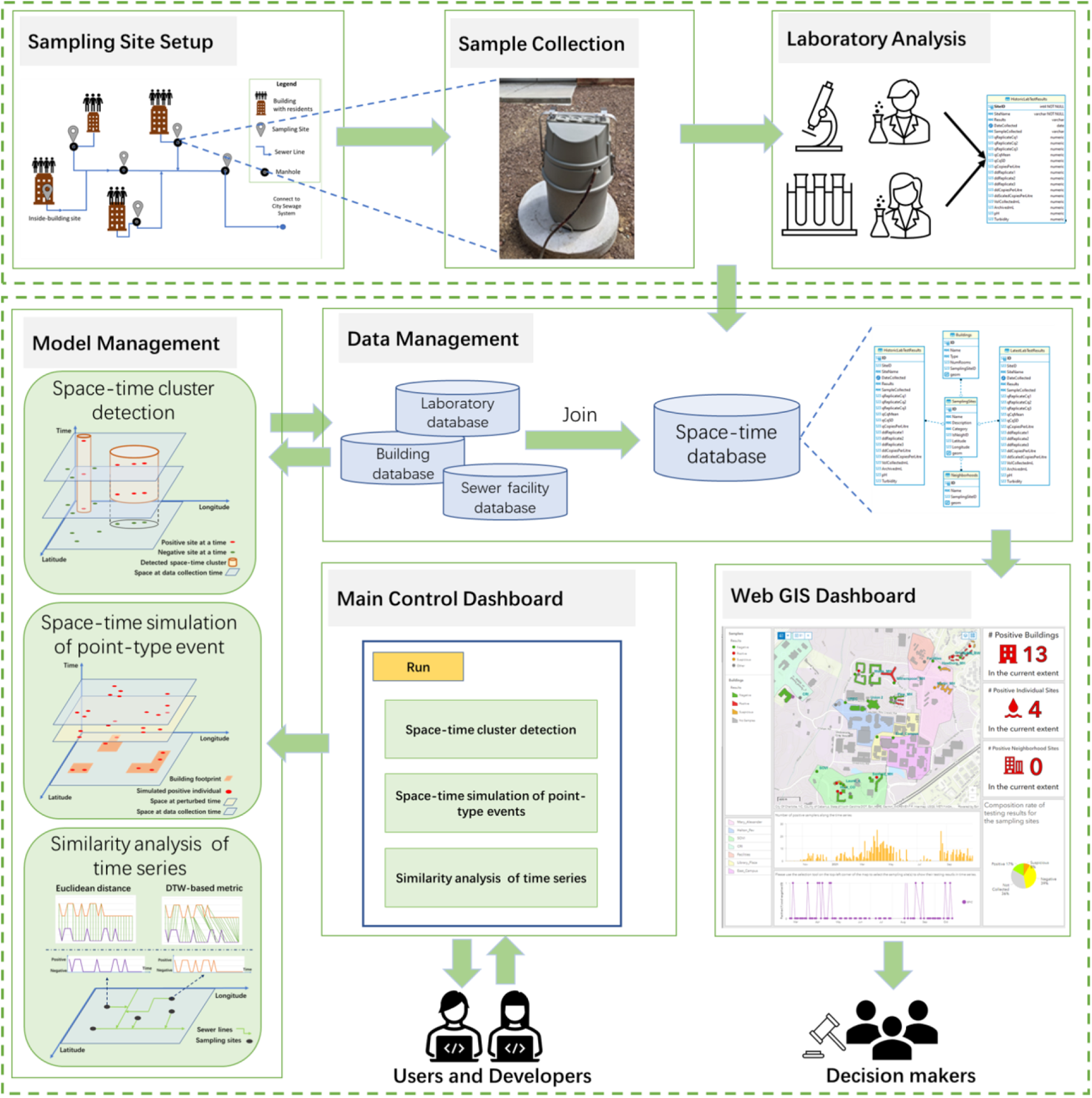
Framework of the web-based spatial decision support system for wastewater surveillance.

Building-level wastewater surveillance typically includes three steps (see Gibas et al., 2021): collection of wastewater samples, sample concentration and RNA extraction, and detection of COVID-19 virus. Various sample-related data are generated from these steps. These data are characterized with space-time stamps and associated with different sampling sites, buildings, and sewersheds. Fundamentally, these data are space-time series that represent various information related to wastewater testing over space and time. Mathematically, our wastewater surveillance data (noted as *W*) can be formulated as in Eq. 1:

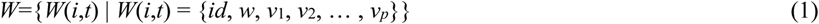

where:

*i*: sampling site ID, *i* ∈ [*1*,*2*,…, *n*];*n*: number of sampling sites;

*t*: ID of time step; *t* ∈ [*t_1_*, *t_2_*,…, *t_m_*]; *t*_1_: beginning date of wastewater sampling; *t_m_*: end date of sampling; *m*: number of sampling dates;

*id*: ID of the sample at site *i* and time *t*.

*w*(*i*,*t*): wastewater testing result for site *i* at time *t* (*w*(*i*,*t*)={0,1}={negative, positive});

*v*_1_(*i*,*t*), *v*_2_(*i*,*t*), …, *v_p_*(*i*,*t*): all other variables associated with site *i* at time *t;* These variables may change over time or not (e.g., testing results will change over time but the ID of associated building(s) will not).

*p*: number of other variables for a sampling site;

Among these variables, the wastewater testing result *w*(*i*,*t*) is a binary variable that indicates whether COVID-19 virus is detected (1: positive; 0: negative) for a sampling site on a specific date. In this study, qPCR detection results from three sample replicates are used to determine whether a sample is considered positive or not. When the virus concentration (mean Cq) values of all three sample replicates are lower (indicating higher viral load) than the empirically determined limit of detection threshold, the corresponding wastewater sample is considered positive. For the purposes of determining administrative response on campus, samples must have all three replicates producing signals to be considered “positive”. Any samples that have only ⅔ replicates producing signals are considered “suspicious” and 1 or fewer replicates producing signals considered negative. This “suspicious” designation is only used for administrative decision purposes. For more detail, please refer to Gibas et al. (2021). In our study here, samples that have 2 or less replicates producing signals are treated as negative (i.e., suspicious and negative samples are merged into a single category: negative).

#### 3.2.1. Spatiotemporal data management and data synchronization

We developed an object-based spatiotemporal data model (see Fig. 4A) to represent spatiotemporally explicit information related to building-level wastewater surveillance for COVID-19 monitoring. Spatiotemporal data models have been developed to represent dynamic geospatial phenomena (Chen et al., 2016; Pelekis et al., 2004; Peuquet & Duan, 1995). Based on spatiotemporal data models, data structures and databases can be designed and implemented to handle data with spatiotemporal stamps. A series of spatiotemporal data models have been proposed in the literature, including snapshot-based, event-based, and object-based (Pelekis et al., 2004). Our spatiotemporal data model is object-based, in which a spatiotemporal object represents a geospatial entity in space and time. As the geometry of sampling sites and buildings does not change, our spatiotemporal data model only needs to take into account change in attributes (non-spatial information) associated with sampling sites or buildings. Thus, a wastewater sample collected at a site at a specific date is abstracted as a spatiotemporal object associated with a set of variables, including sampling site information (geometry: point), building information (geometry: footprint polygon), and lab testing results. Fig. 4B is the entity-relationship (ER) diagram that we used to build the geodatabase based on the spatiotemporal data model. Database tables were created to manage the spatiotemporal data associated with wastewater surveillance (including sampling sites, buildings, sewersheds, historic lab testing results, and latest lab testing results). Further, we used a set of database tables to maintain the relationships between sampling sites and buildings, as well as sampling sites and sewersheds.

**Fig. 4.**
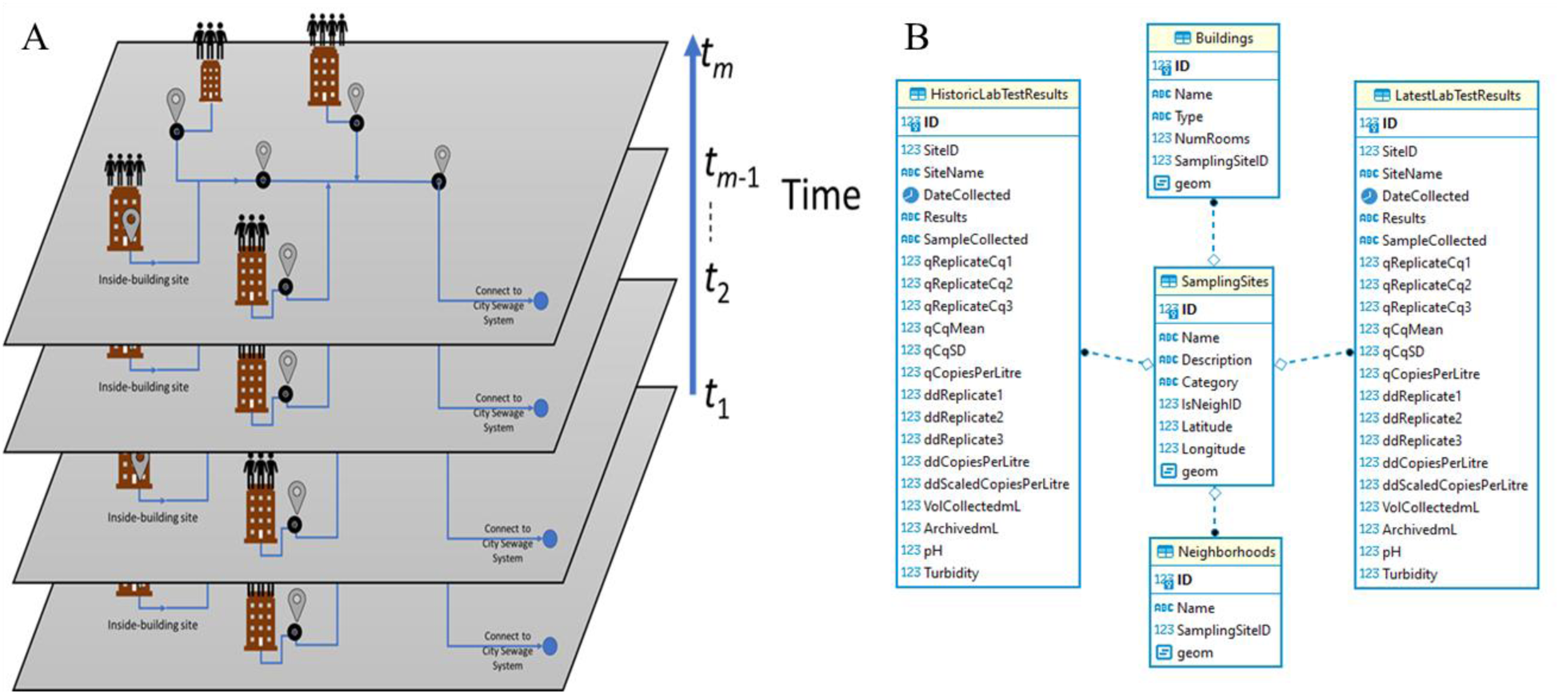
Illustration of spatiotemporal data model (A) and entity-relationship diagram (B) for building-level wastewater surveillance.

We developed an automated synchronization module to upload wastewater testing results once they are available (including real-time and historic data). This automated data synchronization module is implemented within a web-based interface. This synchronization module takes sample testing results (in a delimited file; CSV format) as input and associates these testing results with corresponding buildings or sewersheds (through SQL style left-joins). Then, these testing results are updated to the spatiotemporal database.

#### 3.2.2. Spatiotemporal analysis of wastewater testing results

To address the research questions aforementioned in the Introduction section requires the use of space-time analysis and modeling approaches. We chose to use space-time scan statistics, space-time simulation of asymptomatic individuals, and similarity analysis of space-time series.

##### 3.2.2.1. Space-time scan for cluster detection

In this study, we utilized space-time scan statistics for the detection of space-time clusters of positive wastewater samples reported from wastewater surveillance. We used Kulldorff’s retrospective space-time scan statistic (Kulldorff, 1999; Kulldorff et al., 1998), implemented in SaTScan (version 9.6). A variety of studies have applied the space-time scan statistics approach to detect clusters of covid cases during the COVID-19 pandemic (see, e.g., Desjardins et al., 2020; Hohl et al., 2020; Kim & Castro, 2020; Masrur et al., 2020). However, the space-time cluster detection for COVID-19 monitoring is often applied at large spatial or jurisdictional scales (e.g., state or county level for a country). To our knowledge, this is the first time that the space-time scan statistic is used to detect the presence of COVID19 in wastewater and at a small spatial scale (building level).

The space-time scan statistics uses a cylinder-based scanning window to detect the cluster of space-time objects (e.g., positive wastewater samples here; see Fig.5). The base of the cylinder defines the geographic region covered by the scanning window (the radius of the base is the spatial bandwidth) while the height represents the time duration of the scanning window (i.e., temporal bandwidth). When applying space-time scan statistics, the center of the cylinder is placed at each spatial object (point-types; centroids can be used for polygon-type objects) and the spatiotemporal bandwidth is varied. Then, by using a likelihood ratio test, the number of observed events within and outside the cylinder is compared against their expected values based on Poisson or Bernoulli models (Kulldorff, 1997). Events within a cylinder scanning window with highest likelihood ratio (indication of elevated risk) are identified as a space-time cluster.

**Fig. 5.**
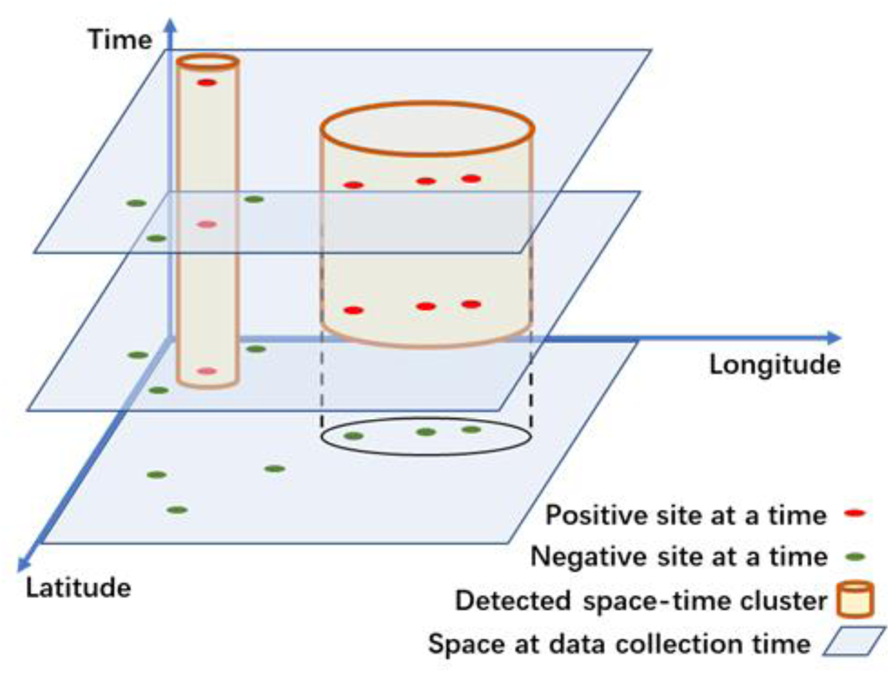
Illustration of using cylindrical scanning windows for space-time scan statistics.

Monte Carlo approach can be used to test the significance of the cluster(s). As the wastewater testing results are a binary variable (positive or negative) in this study, we used the Bernoulli model for the probability model used by the space-time scan statistics.

##### 3.2.2.2. Space-time simulation of asymptomatic individuals

In this study, wastewater testing results from a sampling site are indicative of the situation of the associated building(s)--whether there are presymptomatic individuals in the building. However, the location of the individual(s) within the building is unknown (for privacy protection)—i.e., spatial uncertainty. Further, collected samples on a particular day may be reflective of a prior contamination, keeping in mind that samples were collected every two days or more instead of every day in our study–i.e., temporal uncertainty. Therefore, we used a space-time point pattern simulation approach (see Diggle, 2013) to generate the locations of presymptomatic individuals within the associated building (footprint in polygonal form) and the time that the individuals begin to shed virus. In other words, this approach allows us to simulate space-time locations (where and when) of the presymptomatic individuals, represented as space-time objects in this study.

Fig. 6 illustrates the algorithm of the simulation of space-time point patterns of asymptomatic individuals within buildings. The space-time point pattern simulation begins with footprint polygons of all sampled buildings to determine the spatial location of an asymptomatic individual. A point is randomly generated within the bounding box of the footprint of each building. The point is retained if it is located within the building footprint polygon. Once the spatial location of the presymptomatic individual is determined, the date that the individual begins to shed virus is obtained by randomly perturbing the original sampling date up to *n_perturb* days before (e.g., *n_perturb*=3 in this study). This procedure is applied to each building for a number of Monte Carlo repetitions (e.g., 1,000 repetitions used in this study). After the space-time location is determined, associated sampler site data and testing results are joined. The number of days for perturbation is based on the sampling frequency within a week. For example, 3 days could be used to cover the tri-weekly testing interval. Once simulated results are generated, space-time cluster analysis can be performed on these simulated spatiotemporal point patterns to examine the robustness of space-time clusters detected from observed data.

**Fig. 6.**
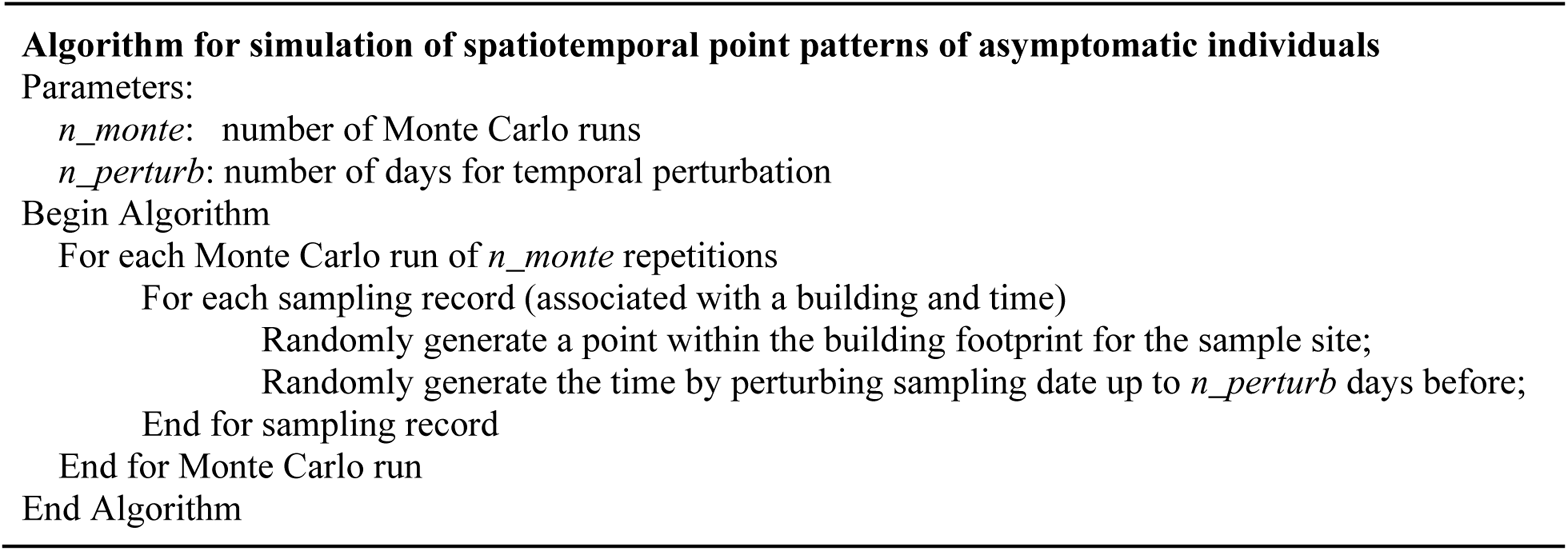
Algorithm of simulation of space-time locations of asymptomatic individuals.

**Fig. 7.**
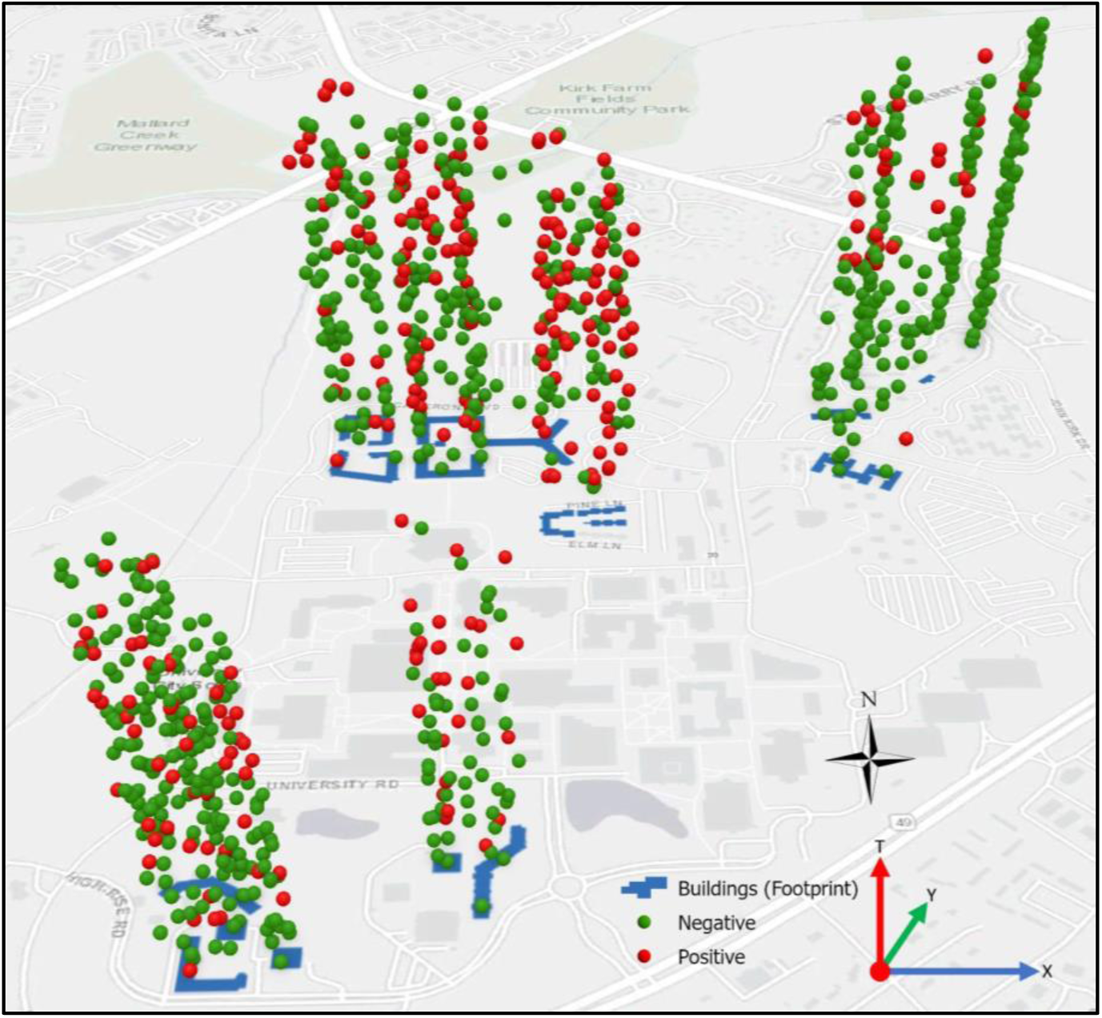
Illustration of a simulated space-time point pattern of asymptomatic individuals (simulated period: January 4^th^, 2021 to May 18^th^, 2021; number of samples: 926; number of positive samples: 264; number of days for perturbation: 3).

##### 3.2.2.3. Similarity analysis of space-time series

To investigate whether any sampling sites show similar responses over time in terms of wastewater testing results, we introduced similarity analysis of time series. We used two similarity metrics, Euclidean distance-based and Dynamic Time Warping (DTW)-based, in this study. Euclidean distance-based metric is a dissimilarity index that evaluates the distance of two time series in the temporal dimension (see Choi et al., 2010). The DTW-based metric allows for comparing time series in terms of shape (see Berndt & Clifford, 1994). DTW is a method that computes the optimal matching between time series (or any sequence patterns) by minimization of distances (Aghabozorgi et al., 2015; Berndt & Clifford, 1994). Given sampling site *i* and *j*, Euclidean distance-based metric (noted as *D_ij_*) between time series of their wastewater testing results can be calculated by Eq. (2). The DTW-based measure (noted as *DTW_ij_*) is represented using the shortest cumulative distance between the beginning and end time steps of wastewater testing results at site *i* and *j* once matching between the two time series is optimized (see Eq. 3).

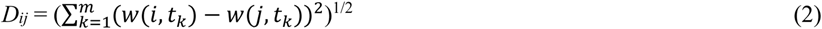

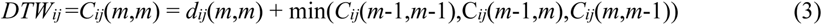

s.t.

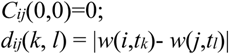

where D*_ij_* and *DTW_ij_* are the Euclidean distance metric and the dynamic time warping metric of the time series between site *i* and *j*. w(*i*,*t_k_*) is the binary testing result of sampling site *i* at time *t_k_*, and w(*j*,*t_l_*) the binary testing result site *j* at time *t_l_* (*k*, *l*∈{1, 2, …, *m*}; *m*: number of sampling dates; defined in Eq. 1). *C_ij_*(*k*,*l*) is the alignment cost between time step *t_k_* of site *i* and time step *t_l_* of site *j*. *d_ij_*(*k*,*l*) is the distance between time step *t_k_* of site *i* and time step *t_l_* of site *j.* |.| is the absolute function that calculates the absolute distance between site *i* and *j*. min(.) is the function to calculate the minimum of costs. The DTW-based measure is derived using a dynamic programming approach (see Sakoe & Chiba, 1978). Each similarity measure is based on the comparison of two time series, which leads to a *n* by *n* matrix of similarity for our wastewater case (*n*: number of sampling sites; see Eq. 1). Once similarity measures are calculated, hierarchical clustering can be applied to these similarity metrics to compare time series of wastewater testing results of all sampling sites.

#### 3.2.3. Web-based mapping and geovisualization

We used a Web GIS approach (Fu & Sun, 2011; see Peng & Tsou, 2003) for the visual presentation of wastewater data and related spatiotemporal analysis results. Based on the spatiotemporal data model, wastewater data are organized in a spatiotemporal database. We publish these spatially explicit data (sampling sites, sewage network, buildings) into geospatial web services that can be mashed up on a client-side web-based dashboard. When new wastewater testing results are available or the previous sample results are updated, the Web GIS module will automatically update these spatiotemporal data (via API) to the client-side web dashboard (including data, charts, and maps). Further, when new sampling sites are added or some existing sites are retired, the Web GIS module allows for updating spatial data and their geospatial web services (e.g., sampling sites in points, sewersheds in polygons).

We used Esri ArcGIS Online (https://www.arcgis.com/) for Web GIS-based dashboard and ArcGIS API for the automated update of wastewater data to the dashboard. Fig. 8 shows the snapshot of our Web GIS dashboard. The web mapping interface shows the locations of buildings, samplers, and sewersheds (aka, neighborhoods), and sewer networks (hidden for confidentiality consideration). Moreover, the color scheme of samplers and buildings indicate the sample testing results (shown in the map legend). Summary of wastewater testing results including number of positive buildings, sampling sites, sewershed sites, and their time series is displayed (for example, in charts). This provides visual and interactive analytics support that can inform decision makers for subsequent decision making on, for example, clinical testing or contract tracing.

**Fig. 8.**
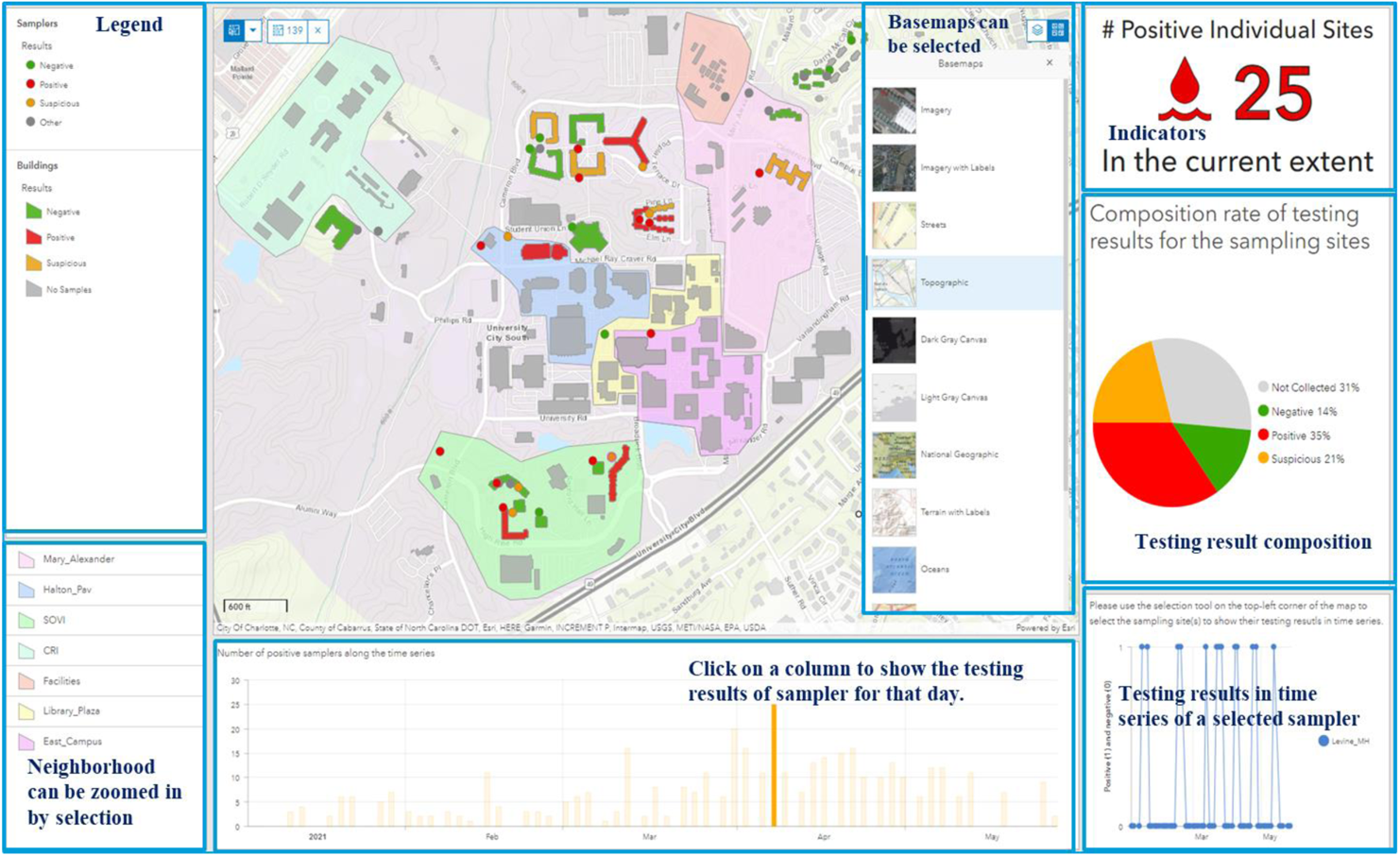
Snapshot of the Web GIS dashboard (sewer networks is hidden due to confidentiality consideration).

#### 3.2.4. Implementation

Our web-based SDSS is implemented within a web server. Jupyter Notebooks (https://jupyter.org/) were used to implement the web-based main interface of the SDSS and access to its individual modules. Table 2 shows the software or libraries used to implement each individual module of the SDSS. We use ArcGIS API for Python to update wastewater testing results to the Web GIS dashboard based on ArcGIS Online. Google OAuth was chosen as the authentication mechanism of a web-based system for automated data synchronization.

**Table 1.** Spatial data collected for the wastewater surveillance work for the study area.

| Spatial Data | Data source | GIS Data Format |
| --- | --- | --- |
| Buildings | Department of Facilities Management of UNC Charlotte | Polygon Vector |
| Sewer lines | Department of Facilities Management of UNC Charlotte | Polyline Vector |
| Sampling sites | Wastewater Surveillance Task Force Group at UNC Charlotte | Point Vector |
| Road network | Department of Facilities Management of UNC Charlotte | Polyline Vector |
| Elevation | U.S. Geological Survey, 3D Elevation Program | Raster |

**Table 2.**
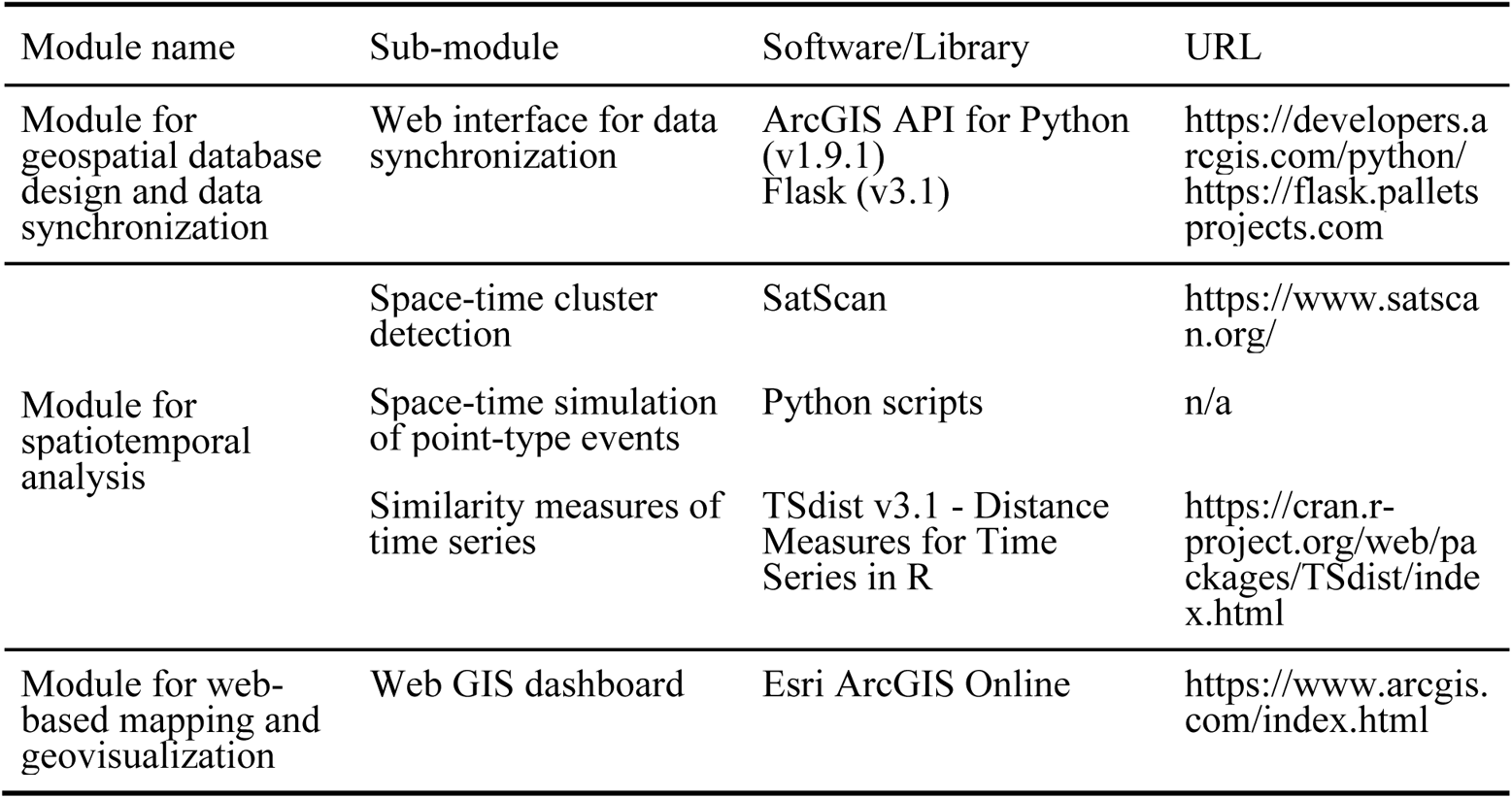
Software or libraries used by the web-based SDSS for wastewater surveillance.

## 4. Results

### 4.1. Overall results

Our wastewater surveillance initiative has been collecting wastewater data since Fall 2020. We have established 38 sampling sites since then. These sites provide strong support for monitoring the COVID-19 situation via the wastewater surveillance approach. Wastewater testing results are uploaded, synchronized, processed, analyzed, and visualized via the web-based SDSS. In this study, we focus on using wastewater testing results from 23 residence hall sites from 01/04/2021to 05/18/2021 (in total 135 days) as we have consistently used these sites to collect samples during this period (results for neighborhoods sites and sampling sites of residence halls that were established or removed during this period were excluded). Sewage samples were collected three times a week on Monday, Wednesday, and Fridays for Spring 2021. This leads to 54 sample collections for each sampling site during the study period (18 weeks times 3 collections per week). However, it is not always possible to collect a sample at every site every time due to variations in flow or unexpected physical obstruction of the autosampler probe. As a result, 926 samples were collected from these 23 sites for Spring 2021. Among them, there are 662 negative (71.49%), and 264 positive (28.50%).

Fig. 9 depicts the number of positive sampling sites during the study period compared to the 7-day averaged number of cases in Mecklenburg County, NC (original data is retrieved from the U.S. Centers for Disease Control and Prevention, https://ephtracking.cdc.gov/DataExplorer/). As we could see, the number of positive sites fluctuates between 0 and 8 before March 24^th^, 2021. After that date, an increasing pattern in terms of the number of positive sampling sites can be observed and lasts for about 2 weeks. This number reaches its maximum (16) on April 9^th^, 2021. After April 9^th^, the number drops to under 10 and tends to show a decreasing pattern over time.

**Fig. 9.**
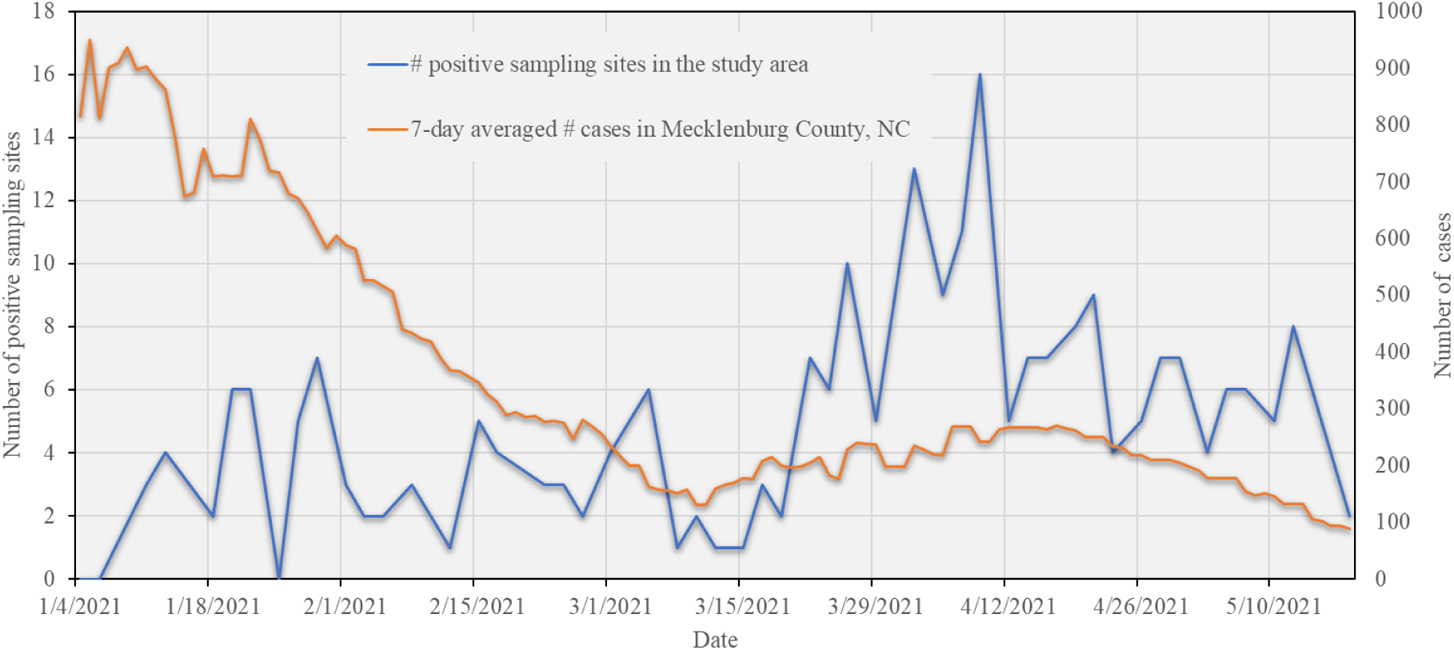
Number of positive sampling sites in the study area and 7-day averaged number of cases in Mecklenburg County, NC over the study period.

The spring semester of the University was postponed to start from January 20^th^, 2021 and Spring Break was changed to the week from February 8^th^ to 13^th^, which was a decision made by the university due to the consideration of the pandemics (number of cases in Mecklenburg County is high in January and February; see Fig. 9). This explains the lower number of positive wastewater samples during the early stage of the semester. An increase in the number of cases in Mecklenburg County (corresponding to the local peak of the SARS-CoV-2 Alpha variant) appeared from mid March to mid April, 2021. Relaxation of local COVID-19 restrictions may also have contributed to this peak (see Executive Orders No. 195 and No. 204 by the North Carolina Governor on February 26^th^ (NC government, 2021a) and March 26^th^ (NC government, 2021b)). This corresponds to (and may explain) a dramatic increase in the number of positive samples on campus during that period. Decreasing trends appeared from mid to late April, 2021 in Mecklenburg County in terms of number of cases and on campus with respect to the number of positive wastewater samples. This can be attributed to the availability of vaccines to more people (increase in vaccination rate). Students started to receive vaccines beginning on March 31^st^, 2021, and vaccines were available to all adults in North Carolina by April 7^th^ (Source: https://governor.nc.gov). Two on-campus vaccine clinics (March 31^st^, 2021, and April 12^th^, 2021) hosted by the university facilitated vaccine uptake by students and faculty. All of these vaccine-related events play an important role in contributing to the decreasing number of positive samples in the final weeks of the semester.

### 4.2. Results of space-time cluster analysis

The use of space-time scan statistics needs to determine the upper limit of the spatiotemporal cluster size bandwidth (spatial bandwidth and temporal bandwidth). For the upper limit of the spatial bandwidth, we set the maximum spatial cluster parameter (corresponding to the percentage of population at risk—i.e., number of collected samples for this study) as 50%. The upper limit of the temporal bandwidth is set to 50% of the duration of the study period.

#### 4.2.1. Space-time cluster analysis results based on samples collected from sampling sites

Fig. 10 and Table 3 depict the space-time scan results based on the collected samples for which the locations of samplers were used as coordinates for space-time scan analysis. Fig. 10 shows the map of detected space-time clusters. One significant cluster (p-value under 5%; based on 999 Monte Carlo runs) was detected that contains two sampling sites lasting from March 17^th^, 2021 to April 30^th^, 2021 (in total 44 days--about 7 weeks). These two sampling sites cover three residence halls. Both the total number of collected samples (population for space-time scan) and number of positive samples (cases) are 34, indicating all collected samples are positive in the detected space-time cluster during these 7 weeks. The detection of this significant cluster is because the three buildings have been used by the University for isolation and quarantine purposes. The relative risk is 3.88 in the detected cluster, indicating the residence halls covered by the sampling sites within the clusters are around 3-4 times higher than those out of the clusters in terms of the ratio of number of positive samples over expected value.

**Fig. 10.**
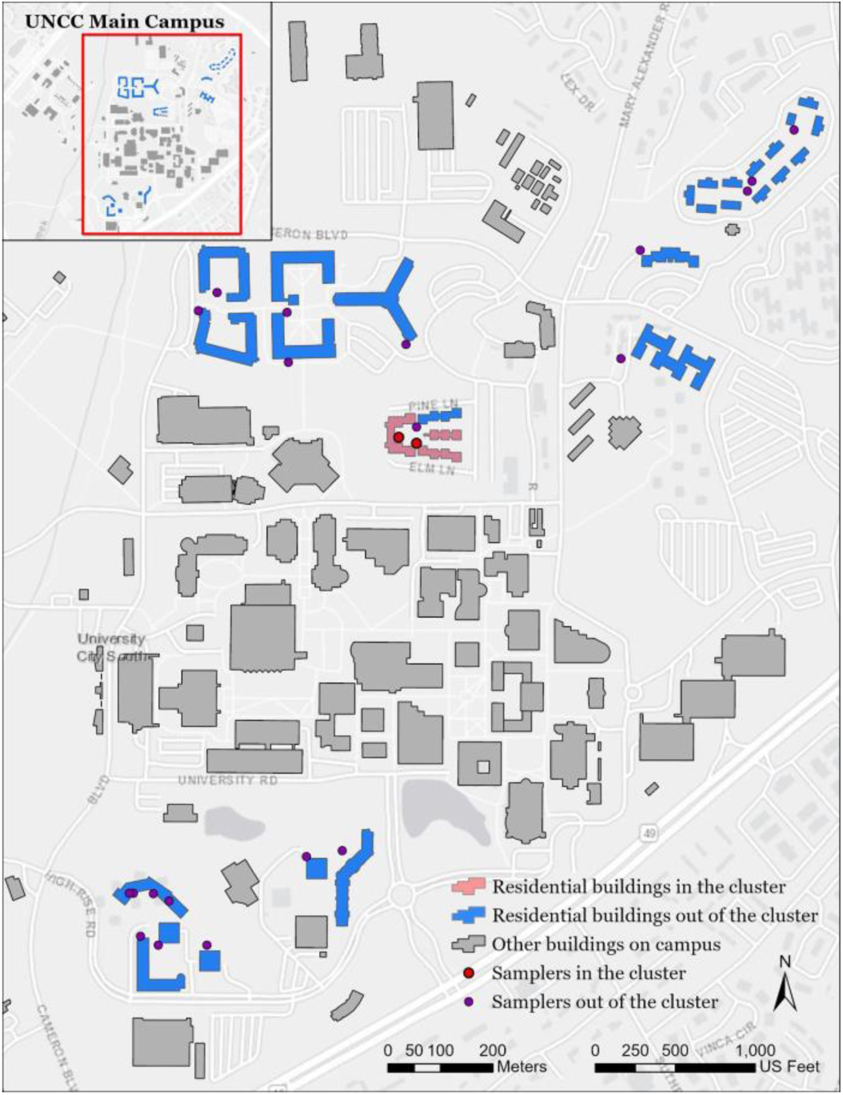
Map of the sampling sites in the detected cluster and corresponding residence halls (sewer networks were hidden due to confidentiality consideration)

**Table 3.** Information of the detected space-time cluster based on locations of sampling sites.

| Parameter | Value | Description |
| --- | --- | --- |
| Time span | 3/17/2021 to 4/30/2021 | Start date and end date of the cluster |
| Population | 34 | Number of collected samples |
| Number of cases | 34 | Number of positive samples |
| Expected cases | 9.69 | The number of samples within the cluster multiplied by the ratio of the total number of positive samples over the total number of samples for the entire study region. |
| Estimated risk | 3.51 | The ratio of the number of positive samples within the cluster over the number of expected cases within the cluster |
| Relative risk | 3.88 | The ratio of the estimated risk within the cluster over that outside of the cluster |
| p-value | 5.2E-15 (p<=0.05) | p-value based on 1,000 Monte Carlo runs |

#### 4.2.2. Space-time cluster analysis results from simulated space-time point patterns

The space-time scan results using locations of collected samples are based on sampling sites. In our case study, these wastewater samples were contributed from individuals living in their residence halls. Our sampling sites are, however, either outside or inside of residence halls, thus posing an issue of locational uncertainty. To address this issue, we used the space-time simulation of point-type events. We associate the binary (positive/negative) wastewater sampling results from sampling sites back to the residence halls. For those sites that cover a single building, once the wastewater testing result from any of these sites is positive, the residence hall will exhibit a positive signal. For a sampling site that covers multiple buildings, all these covered buildings will be positive if the testing results from the site are positive. The number of simulations for generating space-time point patterns was set to 1,000 in this study.

Fig. 11 shows the spatial pattern of residence halls within the detected clusters from 1,000 simulated space-time point patterns (if a simulated presymptomatic individual within a building belongs to a cluster, then we consider the building is within the cluster). There are 8 residence halls that are within significant space-time clusters (at a 95% confidence level). We hide the names of the residence halls for confidentiality purposes. Table 4 summarizes the information of detected clusters based on the 1,000 simulated space-time point patterns. Relative risk within clusters is 2.774, indicating the estimated risk of residence halls within the cluster is 2-3 times higher than that outside the cluster.

**Fig. 11.**
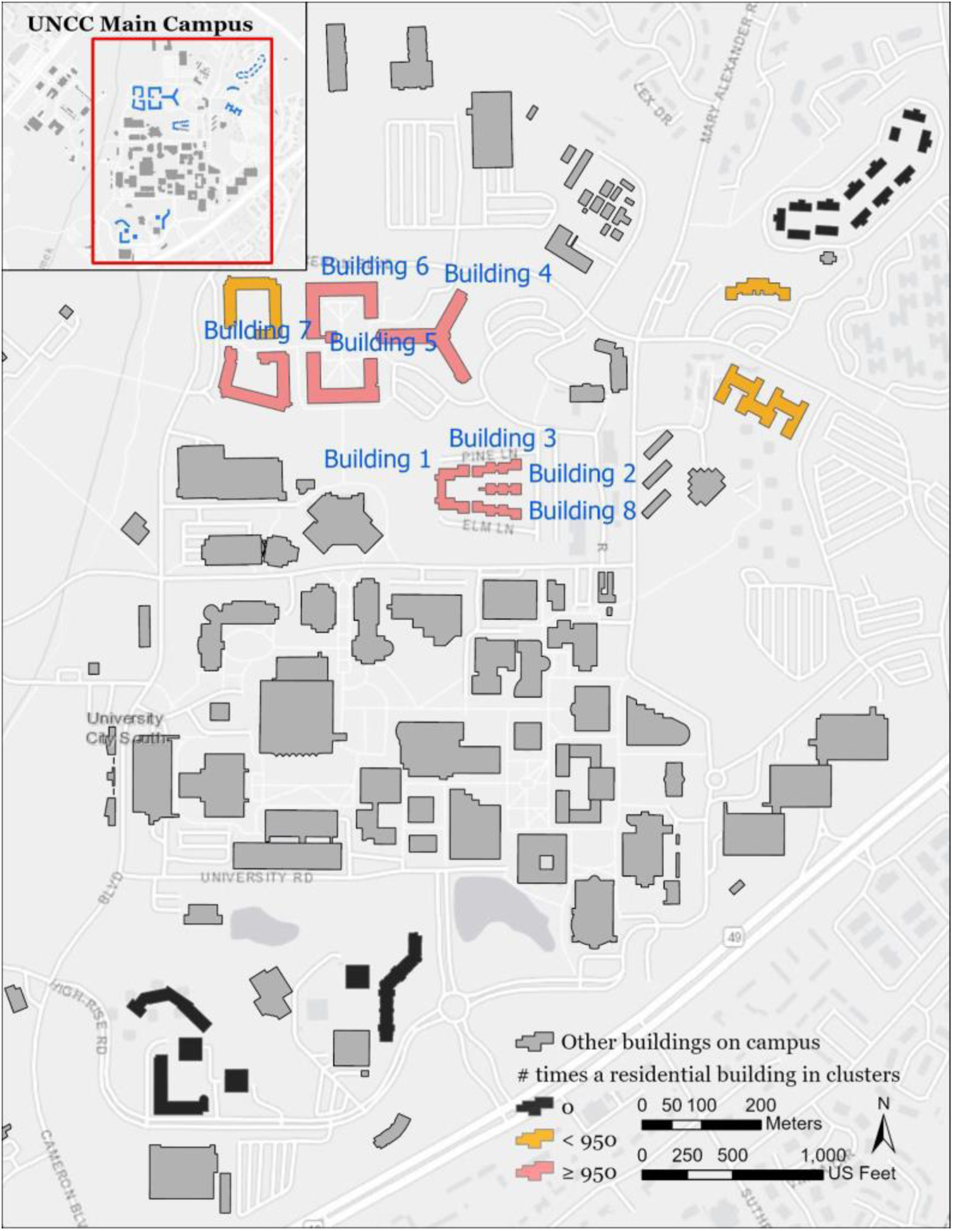
Map of the residence halls in the detected clusters from simulated space-time point patterns (number of simulations: 1,000)

**Table 4.**
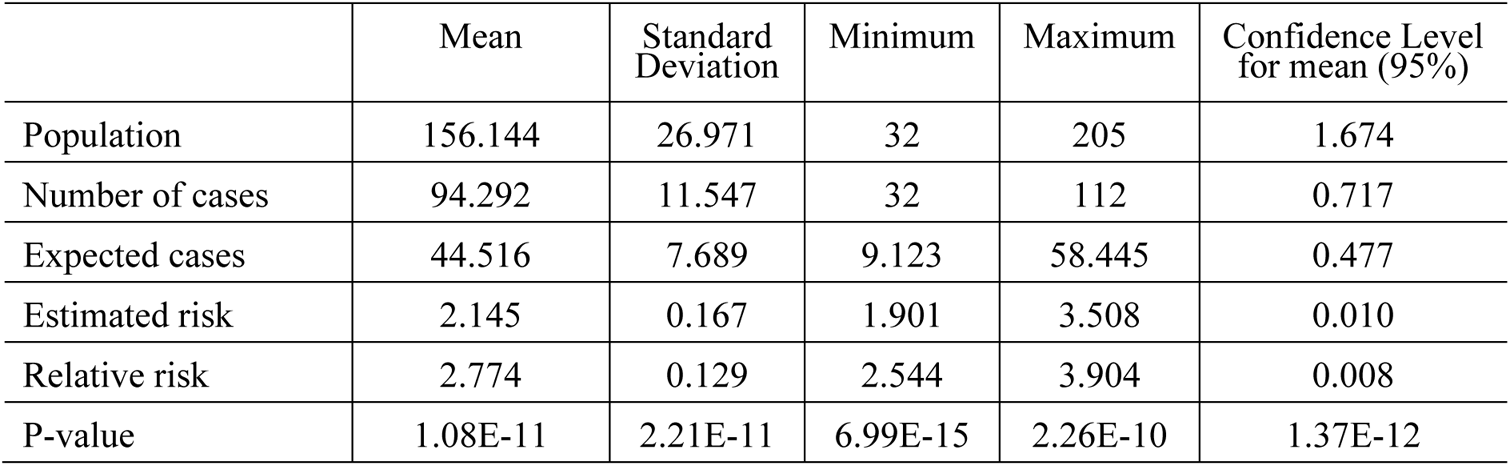
Summary of the clusters detected in 1,000 simulated datasets.

Table 5 depicts start and end dates of each building within clusters, and Table 6 illustrates the number of weeks that the detected space-time clusters from 1,000 simulations last. Fig. 12 shows the histogram of the number of clusters in terms of the start date and end date of a building within detected clusters. It can be observed from Table 6 that detected clusters last from 1 week to 6 weeks, and 92.8% of the clusters last around 3-5 weeks. In general, the significant start date of clusters on each building at high risk concentrates on March 24^th^, 2021 (one exception is March 26^th^ for building 7) and most of them end around April 23^rd^ or 24^th^ (April 20^th^ for building 7), lasting around 1 month. This suggests that the wastewater signals from these 8 buildings correspond to the second peak of the pandemic in Mecklenburg County (see Fig. 9). The three buildings used for isolation and quarantine purposes are included in these 8 buildings.

**Fig. 12.**
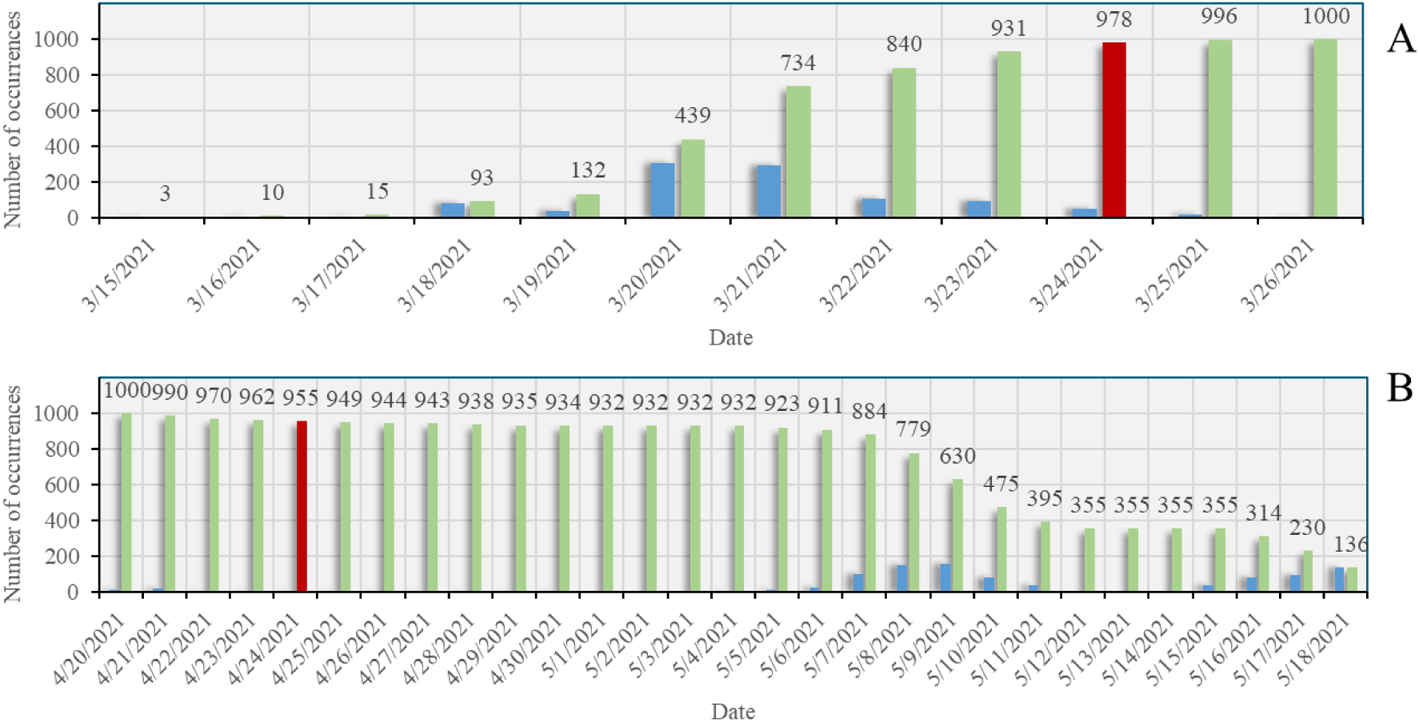
Histograms of the start (A) and end (B) dates that a building (Building 1) was identified as within a cluster (blue) and the number of occurrences that a building was identified as within a cluster over time (green) from simulations. Significant start and end dates (95% confidence level) were colored in red. Number of simulations: 1,000.

**Table 5.** Start and end dates of the buildings detected within clusters based on 1,000 simulations.

| Building index | Start date ($p \leq 0.05$) | End date ($p \leq 0.05$) | Number of days at high risk |
| --- | --- | --- | --- |
| Building 1 | March 23<sup>rd</sup>, 2021 | April 24<sup>th</sup>, 2021 | 33 |
| Building 2 | March 23<sup>rd</sup>, 2021 | April 24<sup>th</sup>, 2021 | 33 |
| Building 3 | March 23<sup>rd</sup>, 2021 | April 24<sup>th</sup>, 2021 | 33 |
| Building 4 | March 23<sup>rd</sup>, 2021 | April 23<sup>rd</sup>, 2021 | 32 |
| Building 5 | March 23<sup>rd</sup>, 2021 | April 23<sup>rd</sup>, 2021 | 32 |
| Building 6 | March 23<sup>rd</sup>, 2021 | April 23<sup>rd</sup>, 2021 | 32 |
| Building 7 | March 26<sup>th</sup>, 2021 | April 20<sup>th</sup>, 2021 | 26 |
| Building 8 | March 23<sup>rd</sup>, 2021 | April 24<sup>th</sup>, 2021 | 33 |

**Table 6.** Number of weeks covered by the space-time clusters detected from simulated patterns (number of simulations: 1,000).

| #Weeks | Frequency |
| --- | --- |
| 1 week | 53 |
| 2 weeks | 18 |
| 3 weeks | 254 |
| 4 weeks | 384 |
| 5 weeks | 290 |
| 6 weeks | 1 |

The use of space-time scan for cluster analysis is computationally demanding because each analysis would need additional 999 Monte Carlo runs for significance testing, and we need to conduct this analysis on 1,000 simulated space-time point patterns of presymptomatic individuals. To address this computational challenge, we deployed these analyses to a high performance computing (HPC) cluster (computing node configuration: dual 24-Core Intel Xeon Gold 6248R CPU with clock rate of 3.00 GHz and 384GB memory). Twenty computing nodes (each with 24 cores--i.e., in total 480 CPUs) were used for acceleration. The parallel computing time of the analysis of a single simulated point pattern on a computing node varies from 7.76 to 16.26 minutes with a mean of 8.42 minutes, while the mean sequential computing time for a single analysis is 139.36 minutes. The total parallel computing time on 480 CPUs for 1,000 analyses is 7.08 hours, compared with the total sequential computing time (on a single CPU) of 2,322.72 hours (around 97 days). As a result, 327.91 times of acceleration was achieved for these analyses by using 480 CPUs.

### 4.3. Results of Similarity Analysis of Time Series

We conducted similarity analyses based on the time series of wastewater testing results from the 23 sampling sites over the study period. Fig. 13 shows the results of similarity analysis with respect to metrics of Euclidean distance and DTW. Both Euclidean distance and DTW are dissimilarity metrics, meaning that the larger the value of the metrics, the more dissimilar the time series of two sites are. We then applied hierarchical clustering analysis to each of the two metrics. Elbow method (Thorndike, 1953) was used to determine the number of clusters based on these metrics. As a result, two clusters were identified with respect to the Euclidean distance metric and three clusters for the DTW metric.

**Fig. 13.**
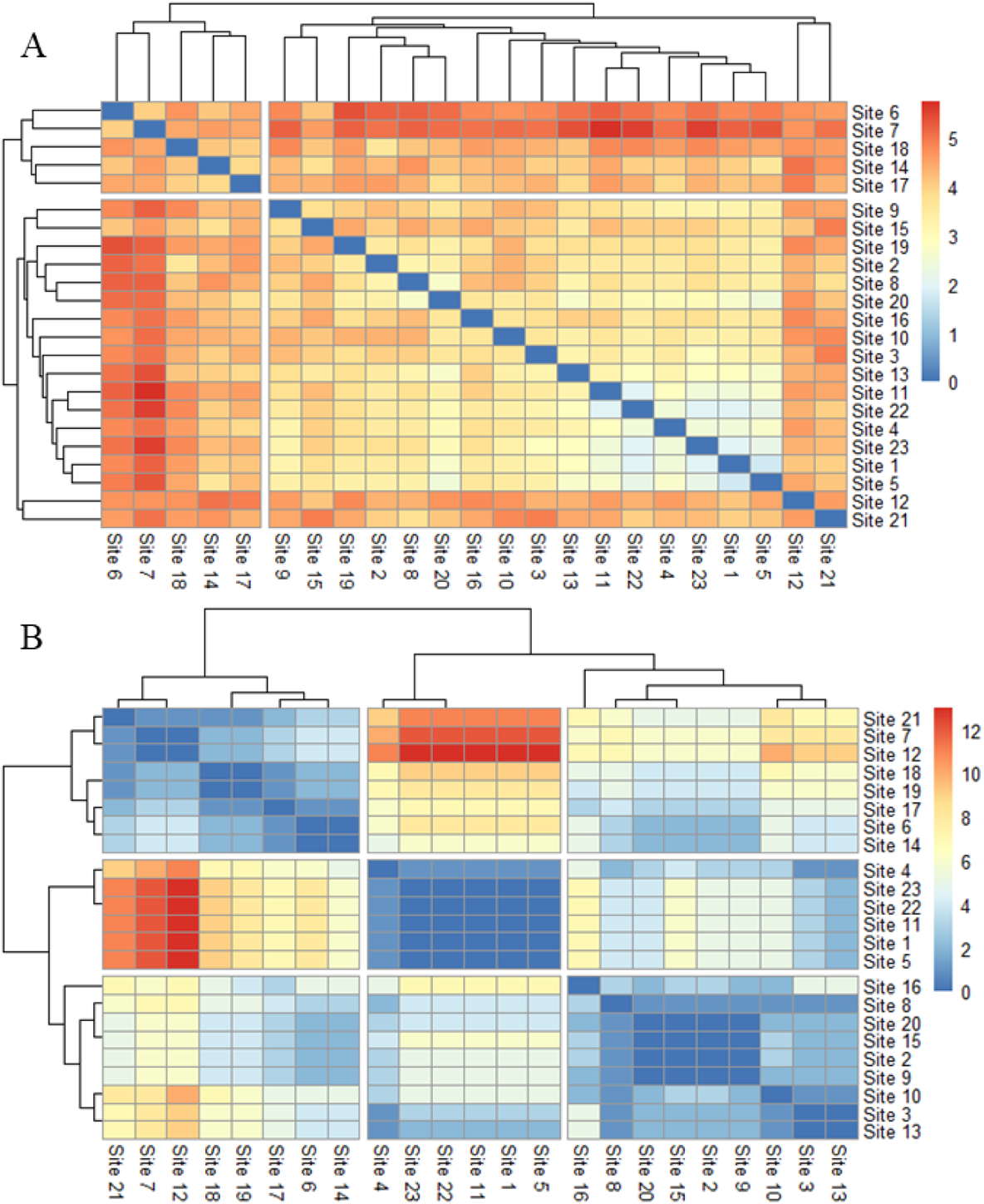
Matrix of similarity metrics. (A: Euclidean distance; B: Dynamic Time Warping).

Fig. 14 depicts the cluster dendrograms of the two similarity metrics as well as the spatial distribution of the identified clusters with respect to each of the similarity metrics. Fig. 15 shows the number of positive sites per week for each group identified by similarity metrics. The Euclidean distance-based metric clusters the sampling sites to two groups, whereas there are three main groups identified by the DTW metric. In terms of Euclidean distance-based metric (see Fig. 15A), group 1 covers five sampling sites, about 22% over 23 sampling sites in this study. The number of positive samples of group 1 fluctuates around 5 positive samples per collection day before and on March 15^th^, 2021. It rises to 10 - 15 positive samples per collection day from late March to mid April and then a decreasing trend appears until mid May. Group 2 has 18 sampling sites, around three times higher than those in group 1. The number of positive samples for each group in the study period tends to be close compared with the total number of sampling sites in each group, indicating that buildings in group 1 are at higher risk of being exposed under virus than those in group 2. We can also observe a rising pattern in the number of positive samples for group 2 in mid March and a decreasing trend from late April to the end of the study period.

**Fig. 14.**
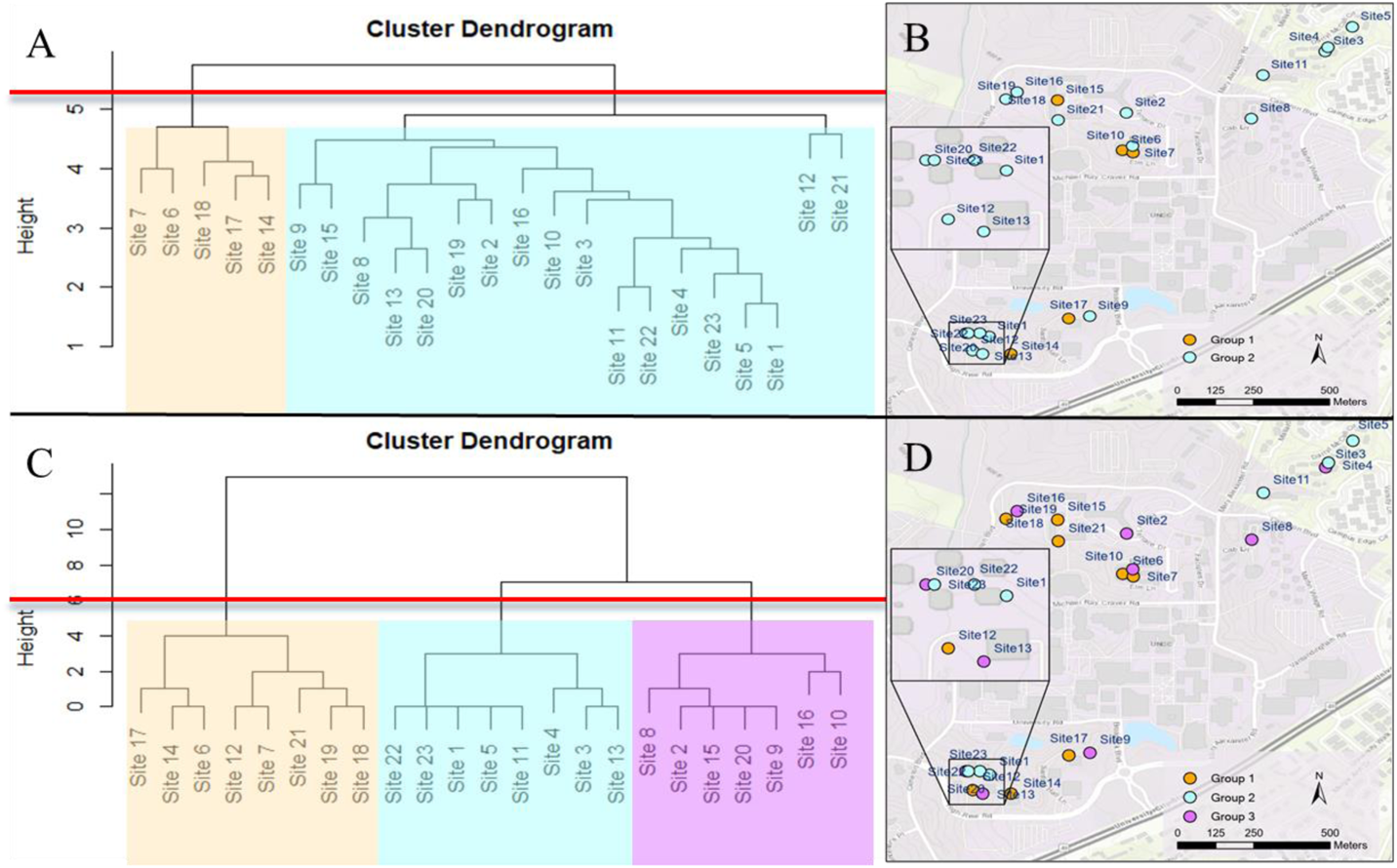
Cluster dendrograms of similarity metrics and spatial patterns of clustered results. A and B are for Euclidean distance metric. C and D are for Dynamic Time Warping metric. The cut-off of the number of clusters (red line) was identified using the Elbow curves. Group 1, 2, and 3 were shaded in orange, blue, and purple.

**Fig. 15.**
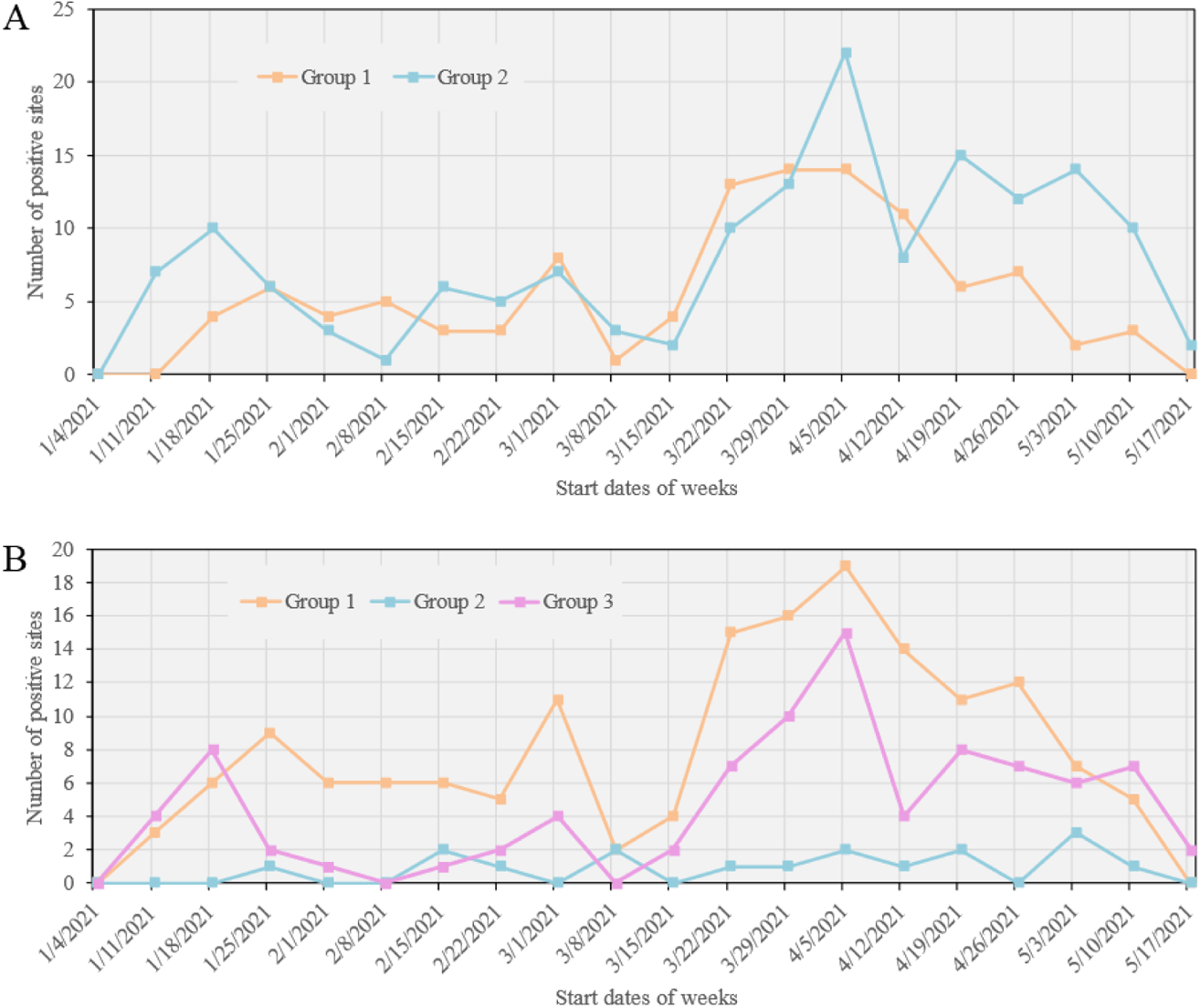
Number of positive sampling sites per week for each group identified by similarity metric over time. A: for the Euclidean distance metric. B: for the Dynamic Time Warping metric. The horizontal axis shows the start date of each week. The last week starting from May 17^th^ only has two-days data available.

With respect to the DTW metric (see Fig. 15B), three groups are identified, where the number of sampling sites are 8, 6, and 9 for group 1, 2, and 3. It is observed that the weekly number of positive samples in group 1 is higher than those of groups 2 and 3 in between January 25^th^, 2021 and May 3^rd^, 2021, covering most of the study period. The number of weekly positive samples in group 2 is higher than that in group 3 especially in the beginning of the study period until February 8^th^, 2021, and from March 15^th^ to May 18^th^, 2021. Group 3 stays between 0 to 3 positive samples per week during this time span. Group 1 and 2 exhibit similar responses to the spread of COVID-19 as we can observe three peaks in the time series: around January 20^th^, March 1^st^, and April 5^th^. Both group 1 and 2 strongly responded to the wave in Mecklenburg County, NC starting from mid March, 2021 (see Fig. 9); however, group 3 did not show a significant reaction to this wave, indicating residence halls in this group appear a relatively lower risk of being exposed to the virus than others during the study period.

Sites 6 and 7 identified in group 1 of both similarity metrics (see Fig. 14) are also detected within the cluster using space-time scan (see Fig. 10), indicating that buildings related to the two sites are more likely to be under exposure of the COVID-19 during the study period. Site 14, 17, and 18 in group 1 for Euclidean distance are also included in the group 1 of DTW metric, indicating these sites also need to be paid attention. Further, group 1 of DTW metric suggests that site 12, 19, and 21 are at relatively higher risk as well. Buildings in group 2 of the Euclidean distance metric appear less likely to be under exposure of the virus than those in group 1. It can be observed that buildings in group 2 and 3 detected by the DTW metric are included in group 2 of the Euclidean distance. Results in Fig. 15B also suggests that the two groups of DTW metric, especially group 2, appear to be characterized by a relatively low number of positive wastewater samples during the study period.

## 5. Discussion

Our web-based SDSS provides support for automating data operations, analysis and modeling, and visualization capabilities within an integrative environment. Wastewater surveillance is dependent on various data that may cut across different spatiotemporal scales. Our web-based SDSS allows for automated synchronization and mapping of these spatiotemporal data. This provides timely support for the early detection of the COVID-19 virus in campus wastewater and thus greatly facilitates the monitoring and mitigation of the pandemic situation in the University. At the same time, the management of space-time wastewater data within this integrated environment can help monitor the status of samplers and their sampling sites. If any issues occur to the autosamplers that lead to the unavailability of samples over time, we could quickly identify and resolve the issues with support from this SDSS, thus ensuring the continual functioning of samplers.

Wastewater surveillance data are spatiotemporally explicit. Spatiotemporal analysis and modeling can be of great help in discovering interesting patterns in these spatiotemporal data, represented by the clusters of positive samples or residence halls detected using space-time scan approach and similarity analysis of space-time series in this study. The combination of the spatiotemporal analysis approaches has been suggested in the literature (see Xu & Beard, 2021). Space-time scan methods, represented by SatScan in this study, allows for detecting the co-occurrence of space-time events (positive samples in this study) within a specific time period (i.e., local- or regional-level analysis). Further, similarity analysis of space-time series offers a means of comparing space-time events over the entire study period--i.e., system-level comparison. Combining these spatiotemporal analysis methods enables us to discover patterns of interest from different levels (with respect to the study system of interest). On the one hand, this combined approach allows for identifying those residence halls where interactions with their residents are at a high risk during specific time periods. On the other hand, it gives us recommendations on the group of residence halls with a lower risk of virus even when it was peaking. This combined analysis approach provides substantial support for addressing spatiotemporal questions (as in the Introduction section). It is also noted that the detection of these space-time clusters may be biased as samples may not be collected from every site each time, which will be investigated in future work. However, in general, these detected clusters from spatiotemporal analysis and modeling provide invaluable and critical support for the University on decisions or guidelines for the prevention of outbreak of the virus and control of virus transmission on campus.

The use of the space-time simulation of presymptomatic individuals was necessary because the relationship between sampling sites and their associated buildings is complicated (instead of one-to-one mapping) and because individuals in residence halls are sources that contribute to the wastewater testing results instead of samplers at sampling sites. The space-time scan results based on simulated individuals in residence halls are different than those based on sampling sites. The former approach detects more residence halls within the clusters of positive wastewater samples. The space-time simulation of the presymptomatic individuals provides an alternative approach for the possible locations of these individuals instead of relying on the sampling sites. While the detected clusters include more residence halls from space-time simulation, it is better than underestimating the number of residence halls that may exhibit strong positive signals of COVID-19 virus in wastewater. Of course, these clusters of positive wastewater samples are based on the space-time scan, which is a statistically based exploratory data analysis approach. The further interpretation of these clusters would require the expert knowledge from the collaboration of domain scientists (e.g., biogenetic professionals), better understanding of the wastewater surveillance system (e.g., sampling sites, residence halls, student interactions), and the incorporation of clinical testing and contact tracing data. In particular, clinical testing data could be used to further improve the space-time simulation of presymptomatic individuals in terms of model calibration and validation. For example, in this study, wastewater samples that have 2 or less replicates producing signals are treated as negative. The use of clinical testing data could help us to fine tune the relationship between wastewater signals and infected individuals for more reliable spatiotemporal cluster analysis.

Web-based GIS is of essence in this web-based SDSS in terms of visual presentation of space-time data related to wastewater surveillance. Web-based GIS technologies and geospatial web services have been increasingly developed and available for the online management and mapping of spatially explicit data. However, the automatic update of data to Web GIS dashboards has been the bottleneck of Web GIS applications. Our web-based GIS and visualization module provides automation support that allows for the automatic update of wastewater sample data to the web GIS dashboard. Specifically, we aimed to reduce the time and number of steps that data are taken from the lab to the dashboard. This module will lead to the saving of tremendous time and cost as required by the update and dissemination of wastewater data that are continuously available over time.

## 6. Conclusions

The web-based SDSS framework presented in this study empowers the management, analytics and sharing of wastewater surveillance-related data at multiple spatiotemporal scales. The SDSS framework serves as a synergistic platform that integrates various types of data based on the spatiotemporal data model. Spatiotemporal analysis and modeling capabilities incorporated in this framework offer a means of unveiling interesting or unexpected patterns from the wastewater data. These patterns may not be easily detected using visual inspection. These data- and model-related capabilities are managed and automated within the SDSS framework to ensure their reusability and the reproducibility of analytic results. This SDSS framework, built in with Web GIS dashboard functionality, will inform critical decision-making and guideline development for monitoring COVID-19 situations in the study region.

Future work of our study includes: 1) integration of 3D BIM-based building model into the web-based environment, 2) adding more spatial modeling capabilities (e.g., spatial simulation for scenario analysis and representation of individual behavior and social behavior using agent-based modeling; spatial optimization for optimal allocation of sampling sites), 3) use of continuous variable of virus concentration in wastewater samples instead of binary indicator for spatiotemporal analysis, and 4) extend the web-based SDSS framework to other or larger regions by, for example, linking to city sewage network and wastewater treatment plants at regional level.

## Data Availability

Data are available from the authors upon request.

## Acknowledgements

The authors would like to thank Chancellor Sharon Gaber, Provost Joan Lorden, and Richard Tankersley, Vice Chancellor for Research and Economic Development and his team for strong institutional support of this wastewater surveillance project, Deborah Thomas, Chair of the Department of Geography and Earth Sciences for facilitating setting up geospatial computing needs for the project. The authors owe thanks to Facilities Management (Greg Cole) and OneIT (Alex Chapin, Elie Saliba) at the University of North Carolina at Charlotte for their support and help on sampling site setup and computing needs. High-performance computing resources used in this project were provided by University Research Computing at the University of North Carolina at Charlotte.

## Appendix

**Appendix 1.** Sources of the information about the University of North Carolina at Charlotte (retrieved year: 2021).

| Sources | URLs |
| --- | --- |
| Faculty and Staff Resources | https://www.charlotte.edu/gateway/faculty-staff |
| Housing and Residence Life | https://housing.charlotte.edu/ |
| University Catalogs | https://catalog.uncc.edu/preview_program.php?catoid=30&poid=8179 |
| Housing and Residence Life | https://housing.charlotte.edu/housing-options/find-your-home |
| Undergraduate Admissions | https://admissions.charlotte.edu/about-unc-charlotte/university-profile |

